# Internal Tremors and Vibration Symptoms Among People with Post-Acute Sequelae of SARS-CoV-2: A narrative review of patient reports

**DOI:** 10.1101/2021.12.03.21267146

**Authors:** Daisy Massey, Anna D Baker, Diana Zicklin Berrent, Nick Güthe, Suzanne Pincus Shidlovsky, Liza Fisher, Connor B Grady, César Caraballo, Richa Sharma, Harlan M Krumholz

## Abstract

To introduce the perspective of patients who have PASC with vibrations and tremors as a prominent component, we leveraged the efforts by [Survivor Corps](https://www.survivorcorps.com/), a grassroots COVID-19 patient advocacy group, to gather information from people in their Facebook group suffering from vibrations and tremors. Survivor Corps collected 140 emails and 450 Facebook comments from members. From the emails, we identified 22 themes and 7 broader domains based on common coding techniques for qualitative data and the constant comparative method of qualitative data analysis. Facebook comments were analyzed using Word Clouds to visualize frequency of terms. The respondents’ emails reflected 7 domains that formed the basis of characterizing their experience with vibrations and tremors. These domains were: (1) symptom experience, description, and anatomic location; (2) initial symptom onset; (3) symptom timing; (4) symptom triggers or alleviators; (5) change from baseline health status; (6) experience with medical establishment; and (7) impact on people’s lives and livelihood. There were 22 themes total, each corresponding to one of the broader domains. The Facebook comments Word Cloud revealed that the 10 most common words used in comments were: tremors (64), covid (55), pain (51), vibrations (43), months (36), burning (29), feet (24), hands (22), legs (21), back (20). Overall, these patient narratives described intense suffering, and there is still no diagnosis or treatment available.

## Introduction

Post-Acute Sequelae of SARS-CoV-2 infection (PASC), also known as long Covid, is a condition that is marked by protean manifestation that vary considerably among individuals.^1-4^ The heterogeneity of PASC requires attention to specific clusters of individuals who are suffering from similar symptoms. Proper clustering may enable efforts to identify biological signatures that help elucidate underlying mechanisms and can guide the development of diagnostics and therapeutic tools and strategies.

Some people with long Covid manifest symptoms that they describe as vibrations and tremors. To date, these symptoms, as experienced by patients, have not yet been well described. There are reports of patients with myoclonus, but they are described from the perspective of clinicians.^5-12^ No underlying cause of the myoclonus episodes among patients with PASC has been identified.

To introduce the perspective of patients who have PASC with vibrations and tremors as a prominent component, we leveraged the efforts by [Survivor Corps](https://www.survivorcorps.com/), a grassroots COVID-19 patient advocacy group, to gather information from people in their Facebook group suffering from vibrations and tremors.^13-15^ We conducted a qualitative analysis of the responses and organized them according to prominent themes. The goal of this study was to give voice to the patient perspective and identify a potentially important cluster of similar symptoms that people with PASC are reporting.

## Methods

### Study Design and Sample

The idea for the study originated with the experience of Heidi Ferrer and the initiative of her husband, Nick Güthe. Heidi had severe manifestations of the sensations of vibrations and exhibiting of tremors with onset early in the pandemic. She ultimately committed suicide as she found the symptoms intolerable.

The qualitative approach was chosen because few studies have investigated the patient perspective on these symptoms. Qualitative research is particularly well suited for exploratory studies for which previous literature is limited.^16^ Such studies are useful for generating hypotheses that can later be tested with quantitative data and analyses.^16,17^ In this case, the method was modified to be used with data generated by patients in response to a call for information. This study is considered a starting point for further research.

Nick Güthe spoke with Survivor Corps, who initiated a call for people in their Facebook group of ∼180,000 to report their experience with vibrations and tremors among those with PASC (posts can be found in Supplement 1). This Facebook group represents people who joined and there is no information about their characteristics or how many of them have had an infection with SARS-CoV-2. It is a convenience sample.

Survivor Corps shared the responses for the purpose of a qualitative study with the objective of elucidating themes in the responses and providing an opportunity to give voice to the patient experience. The study sample represents a distillation of the responses from the 140 people who sent information. This study received exemption from the Yale IRB.

## Data Collection

The data were unstructured responses to Survivor Corps’ call for information. There was no secondary follow-up for more information than what was initially provided. There were no structured prompts beyond an initial call for information from people with PASC suffering vibrations and tremors. The responses were aggregated for data analysis.

### Emails

The patient group, Survivor Corps, collected and deidentified emails from people experiencing vibrations or tremors. The group requested emails from members via Facebook and a newsletter (Supplement 1). Emails were received between July 15, 2021 and July 27, 2021.

### Facebook Comments

Facebook comments on the Survivor Corps Facebook page were also analyzed.

First, we analyzed the comments on one member’s post that was aimed at soliciting Facebook messages from people experiencing tremor or internal vibration symptoms. The following was posted on July 14, 2021:

> “Hi, to anyone on this group. It’s Nick Güthe, Heidi Ferrer’s husband. A study is forming with a top doctor for Long Haulers with Neurological tremors similar to Heidi Ferrer’s -- Tremors or internal vibrations. If you have these symptoms and want to be included please comment below. This isn’t a clinical trial but an attempt to gather data and stories to help get funding, bring attention to these symptoms which are so destructive to any Long Haulers physical and mental health.”

Facebook comments in response to this post were analyzed on July 16, 2021. Comments were coded by a clinician analyst (CC) as reporting symptoms, unrelated to symptoms, possibly related to symptoms. All posts coded as reporting symptoms or possibly related to symptoms were included in subsequent analysis.

Next, we analyzed the comments responding to a poll posted in the group to understand members’ symptoms. The poll was posted on June 27, 2021, and was titled “Vibration/Buzzing/Pain Poll.” There were 20 answer choices, each a statement relevant to vibration, tingling, buzzing, and neuropathic sensations (Supplemental Table 1). The analysis included only the comments posted in response to the poll.

## Data Analysis

### Emails

Exact dates and names were removed and replaced with month and year and five-year age brackets to protect anonymity. Email data were analyzed using common coding techniques for qualitative data and the constant comparative method of qualitative data analysis.^18^ Coding of the data was accomplished in iterative steps. An initial code list was generated after an initial evaluation of the data by members of the research team (HMK and DM). During its development, the code structure was reviewed by the full research team for logic and breadth. A total of 7 broad themes were ultimately developed and served as the basis for final text review and organization of the data.

Using this final version of the code structure, members of the research team (HMK and DM) independently coded all transcripts, then met as a group to code in several joint sessions, achieving consensus and assigning codes to observations by a negotiated, group process. Coded data were entered into a software package designed to handle unstructured qualitative data (nVIVO version 12) to assist in reporting recurrent themes, links among the themes, and supporting quotations.

### Facebook Comments

Facebook comments were shorter and contained less narrative than emails, so we instead used Word Cloud methodology to visualize the prevalence of terms using the *quanteda* [1], *wordcloud* [2], and *tm* [3] packages in R v4.0.3 (R Development Core Team, Vienna, Austria) [4]. We set the minimum frequency of words to be included into the Word Cloud to 3, the maximum number of words to 200, and removed extraneous filler words.

## Results

### Patient Themes

After review of the email data, respondents’ comments reflected 7 domains that formed the basis of characterizing their experience with vibrations and tremors. These domains were: (1) symptom experience, description, and anatomic location; (2) initial symptom onset; (3) symptom timing; (4) symptom triggers or alleviators; (5) change from baseline health status; (6) experience with medical establishment; and (7) impact on people’s lives and livelihood. There were 22 themes total, each corresponding to one of the broader domains. Sample quotations are listed to illustrate the themes.

1) Symptom Experience, Description, and Anatomic Location

Theme 1. Vibrations and tremors were described concomitantly, with descriptions of internal vibrations, visible tremors, and some people experiencing both.

> *Example 1*.
>
> “Sometimes my entire body feels like it’s humming and trembling. It’s like I’m sitting on a huge speaker with the volume all the way up. Through the progression of the last few months, the complete body humming has slowed down, but still happens 5-8 times a month. My hands and legs also began tremoring about the same time as the whole body. My legs bop up and down aggressively at times. I’m not cold, I’m not restless, but my legs visibly move up and down like I’m tapping my foot. My hands have been the worst of it. I felt like they had improved a few months ago, but they’re back with a vengeance. I’m not hungry ever. I know I need to eat, and my go-to has been soup. By the time I get the spoon 4 inches above the bowl, and close to my mouth, all of the broth has been “shaken” off. My handwriting.. awful. Sometimes I absolutely cannot stand myself and just go to bed. That happens more often than not. I usually try to hide my hands in my pockets or under the table, but am not always able to do that. When my whole body is tremoring, I find it a lot more difficult to focus and to get anything accomplished.”
>
> *Example 2*.
>
> “Internal vibrations started about 3 weeks after. They started in my back and back of upper thighs. It felt like I was sitting on a vibration massage chair. They never went away but would vary in intensity. February 2021 I started having restless left arm at bedtime where my left arm would flap until I fell asleep. On [May 2021] it progressed to full body myoclonic movements lasting up to 30 minutes.”

Theme 2. Vibration or tremor site varied, from the entire body to localization in extremities, chest, abdomen, and other locations.

> *Example 1*.
>
> “I experience daily internal tremors/vibrations all over but mainly inside my brain and chest. I have external tremors in my legs, arms and chest.”
>
> *Example 2*.
>
> “Still suffering with symptoms. One of which is tremors and internal vibrations primarily in the legs and feet but do sometimes occur in arms.”

Theme 3. Vibrations and tremors occurred with other symptoms of varying number.

> *Example 1*.
>
> “I also experience relentless headaches. I have had the same one, in varying degrees, since October. Crushing fatigue. Vivid dreams. And the worst, word retrieval.”
>
> *Example 2*.
>
> “Here is a list of my current symptoms that I have over a year after my acute infection:
>
> 1. Extreme Fatigue 2. Exercise Intolerance/Post-Exertion Malaise (Physical and Mental) 3. Short-Term Memory Loss. Must carry a notebook to remember things 4. Brain Fog 5. Muscle Weakness 6. Dizziness. Can’t Drive 7. Seizures 8. Headaches/Migraines 9. High Blood Pressure 10. Cold Hands and Feet 11. Ringing in Ears Tinnitus 12. Hoarseness/Loss of Voice 13. Loss of Coordination in Hands 14. Burning Sensation on Skin Lower Torso”

Theme 4. Vibrations and tremors could cause severe pain.

> *Example 1*.
>
> “My brain shakes after a few hours inside my head, my face starts to tingle and numb, and then the full head shaking seizures start. I have severe head pain and nausea constantly from all the seizures.”
>
> *Example 2*.
>
> “That week of unrelieved spasms lee my body barely able to move. Like paralyzed. I had 3 natural child births. I could not fake such 10/10 pain. I have never felt such intense pain, I thought my back would break and my right arm would be completely dislocated twisted out of socket. I could not breathe at times due to the Laryngeal spasms and diaphragm spasms.”

2) Initial Symptom Onset

Theme 5. Vibration and tremor initial onset varied, from the day of initial infection to weeks or even months later.

> *Example 1*.
>
> “Symptoms started on [July]. About 3 weeks later, I developed tremors. I’ve had them ever since.”
>
> *Example 2*.
>
> “I contracted COVID from an ICU patient in May 2020. A few days later terrible headaches, loss smell, lung, cardio, eye damage. Ongoing problems fatigue, Headaches, Migraines, Imbalance, Dizzy, Vertigo, SOB, COPD, Brady/Tachycardia, SVT, chest pain, Gastric, swallow, voice, cognitive, exercise intolerance, several leaves of absences off work. I had many scans, tests, labs with both normal results and damage results. I have all Records. Then few months later unrelenting Neuro issues vibrations, ripples, tremors, became intense foot cramps, painful ankle, foot drop, leg spasms started mostly r foot. I had to wear ankle brace use cane. Very difficult to sleep. Husband could see ripples under skin and feel the vibrations at times. Body is constantly “on”, pain, numb, burning, briar patch, walking on nails, spikes r foot, can’t put r foot flat. Also forearms, hands, r side worse.”

Theme 6. Vibrations and tremors occurred following or during acute COVID-19 infections that varied from mild to severe.

> *Example 1*.
>
> “I was diagnosed with COVID-19 in July of 2020. I spent 2 months in the ICU and 9 days on ECMO.”
>
> *Example 2*.
>
> “I had a moderate case of Covid in November of 2020 with multiple symptoms. I was never hospitalized. I was recovered one week before my first Long Covid symptom of shortness of breath began.”

3) Symptom Timing

Theme 7. Vibration and tremor episodes could be brief, or could be prolonged, even constant.

> *Example 1*.
>
> “The internal tremors in my chest generally only last for about 5 seconds or so and then completely subside. The ones in my abdomen are more rare and have lasted for longer, but still less than a minute or so in general.”
>
> *Example 2*.
>
> “Now here at almost 8 months post covid, I have dealt with these horrible tremors daily. They are constant, they don’t come and go. They are 24/7. I feel them more when I am still and resting or at night and early morning, or during naps. If I can just get up and get going most days, I don’t notice them much, unless the intensity increases and I get breakthrough pain or headaches. But the night time always reminds me they are still there.”

Theme 8. Vibration and tremor episodes could occur constantly, daily, or only when relapses occurred.

> *Example 1*.
>
> “The tremors and the dizziness are daily challenges.”
>
> *Example 2*.
>
> “Tremors and ‘vibrations’ are a few of the many ongoing symptoms. The tremors I notice in my hands and toes. It lasts for about 10 seconds or less about every 2 to 3 days. The vibrations I notice when I first lay down for bed at night. It lasts about seconds.”

Theme 9. Vibration and tremor symptoms could completely resolve temporarily and could return up to months later.

> *Example 1*.
>
> “Even now, almost eight months out in July, I still occasionally experience these. They do not occur daily, but do generally happen in conjunction with the relapse of other symptoms, such as mild chest, throat, and back pressure/tightness, and tingling in my extremities.”
>
> *Example 2*.
>
> “Sometimes my entire body feels like it’s humming and trembling. It’s like I’m sitting on a huge speaker with the volume all the way up. Through the progression of the last few months, the complete body humming has slowed down, but still happens 5-8 times a month. My hands and legs also began tremoring about the same time as the whole body. My legs bop up and down aggressively at times. I’m not cold, I’m not restless, but my legs visibly move up and down like I’m tapping my foot. My hands have been the worst of it. I felt like they had improved a few months ago, but they’re back with a vengeance.”

Theme 10. People experienced vibrations and tremors over different time periods (even if they were episodic), and some did not have improvement in symptoms after more than a year.

> *Example 1*.
>
> “I got tremors on [December 2020] the day I got covid & it increased to where I had it in my whole body vibrating on head as well increasing during activity. It’s been very debilitating & frustrating. I have been a patient in Rochester Minnesota Mayo Covid Clinic and my symptoms have lessened. However, it is now it is [July] and the tremors have not subsided.”
>
> *Example 2*.
>
> “I had Covid in early February 2020. I did not know I had it. Severe Headache, sore throat, rash and crushing fatigue. Got better in a flash. 5 months later, the floor dropped out. By early October, hand tremors started. They have diminished and come back intermittently. It is now 18 Months after the initial infection.”

4) Symptom triggers or alleviators

Theme 11. Exercise and activity were associated for some with onsets of tremors and vibrations.

> *Example 1*.
>
> “Since then I notice that if I get my heart rate up too high (which could be anything above 110-15) the tremors and vibrations are made worse.”
>
> *Example 2*.
>
> “I have had long covid for 6 months now, and I get tremors/vibrations/buzzing nerves whenever I overextend myself.”

Theme 12. A variety of self-treatment strategies, such as diet modifications and humming, were used to alleviate tremors and vibrations.

> *Example 1*.
>
> “Regarding the nerve issues, I have found very recently that humming in the morning helps me to stop the vibrations faster. I suspect our vagus nerve is being affected.”
>
> *Example 2*.
>
> “I have experimented with supplements, removing medicines, and diet. Removing all sugar and processed foods from my diet has reduced the internal vibrations. If I ever slip up, the intensity is extreme.”

5) Change from Baseline Health Status

Theme 13. People with vibrations and tremors had varying health states before their COVID-19 infection, from those who were completely healthy to those with pre-existing conditions.

> *Example 1*.
>
> “I was a healthy [30-35-year-old] marathon runner. Now I’m a [30-35-year-old] individual who is grieving who the person was, figuring out how this new body works, realizing it still works differently day-to-day, while also having physicians refuse to treat me.”
>
> *Example 2*.
>
> “I am an extremely physically active [50-55-year-old] peri-menopausal Canadian female with no pre-existing conditions except for being a migraine sufferer all my adult life.”

6) Experience with Medical Establishment

Theme 14. Medical testing failed to reveal the mechanism of either tremors or vibrations.

> *Example 1*.
>
> “Had a brain CT & brain MRI (all normal). 2. After a short duration of sleep or a nap (15-30 minutes), upon waking I feel that my heart is racing, like palpitations. I feel shaky…as if there is a fast motor running inside me. I’ve now learned to sit up slowly and give it a few minutes and then it goes away. Diagnostic cardiac/pulmonary tests I’ve had: EKGs, CT Thoracic Stress test, Echocardiogram, Pulmonary Function & Blood Oxygen Stress (all normal). Had over 50 lab/blood tests and all normal.”
>
> *Example 2*.
>
> “Had MRI that showed micro clots and white matter. Had an EEG but do not know results.”

Theme 15. Tremor and vibration symptoms were sometimes doubted or dismissed by doctors.

> *Example 1*.
>
> “Some doctors have been very dismissive and charted that ‘she just prefers not to walk’. When I arrived at ER, my previous Neurology MD requested Pysch consult..this delayed medical evaluation. They did not believe my pain nor that I could not breathe as my husband begged them to roll me to my side as they tied me flat to the bed. Most of the time I could barely speak as spasms affected my mouth. The previous Neurologist told me to ‘dumb myself down as a nurse and quit researching and causing myself stress’.”
>
> *Example 2*.
>
> “In August 2020, when my heart started racing for hours, I tried to speak to my pulmonologist, and they had a local doctor call who was running a test site, and wanted to assure me it wasn’t possible for me to have Covid because (1) it wasn’t in our area yet, but I was in Boston the day before they announced an outbreak, (2) that symptoms only last 2 weeks max, and I explained Long Covid. (3) That I shouldn’t read research that I can’t understand, but I used to work in a med related field, trained in pre-med/vet including epidemiology, was part of a 2010-2015 pandemic task force related group, and currently work in genetics research, so then she said that (4) this is all in my head and I just need to get over myself and get back to work full time (I was) and that work makes people better (fatigue says no). She yelled at me for over 2 hours. She now is in charge of home bound patient care and tells people they need to have compassion for Long Covid as it can take ‘a few weeks to recover’ and then tells stories about people who recover quickly being examples of ‘good people’ while insinuating that those who stay ill don’t want to heal. My doctors have run the gamut of not believing me; to finally agreeing that Long Covid is real but that since I don’t have a positive test, I can’t have it; to even if I have Long Covid, they don’t have treatment, so short of an ER admit, I just need to suck it up.”

Theme 16. The vaccine was associated with both improvement in symptoms for some people, and a relapse in symptoms for others.

> *Example 1*.
>
> “I had slight tremors in my hands after originally getting sick in June 2020 and after my second vaccine, on [May 2021] (two days later) I started having more seizure-like symptoms.”
>
> *Example 2*.
>
> “I had the internal vibrations intensely during my year of long haul. Since the vaccine most of my symptoms have abated or significantly diminished. I do still have some of the internal vibration though, especially after exertion.”

Theme 17. Medications have been provided for tremor symptoms, with varying results.

> *Example 1*.
>
> “They tried Gabapentin but it didn’t stop them. From March through [July] I had 5-9 seizures trying to fall asleep every night, but they only happened at night. Since the Gabapentni wasn’t working they switched me to Topamax. I was ramping into Topamax throughout June but it wasn’t working either. My family and I got sick again at the end of June with something viral (multiple negative Covid, Flu, and Strep tests that week). The same time they adjusted my Topamax dosage up again. Something changed again.
>
> Suddenly I was having non-stop seizures back to back and was hospitalized [July] and also again on [July] at two different hospitals for uncontrolled seizures. My EEG showed normal and they switched meds again to Keppra. So far I am on 500 mg Keppra 2x day and ramping up, but my seizures are uncontrolled. The meds work only for a few hours and I have had to go on Short Term Disability from work.”
>
> *Example 2*.
>
> “He also put me on the very lowest dose gabapentin 3x per day. I began the medication the same night. The next day I had a terrible headache but something felt different. I continued the meds 3x per day like directed. After 1 week I began to be able to get up and move a little. I began doing dishes and light house work. My family was rejoicing. I was improving. After a few weeks of being on the medication I could tell it was helping with all the pain. The tremors were still there, but were farther in the background, if that makes sense?.. as if they had been put on soft mute. The meds weren’t stopping the tremors but calmed them I guess.”

7) Impact on People’s Lives and Livelihood

Theme 18. Vibrations and tremors were associated with mental health effects, including anxiety, depression, and suicidal thoughts.

> *Example 1*.
>
> “The psychologist who saw me for 30 minutes gaslit me saying I need to exercise more as my severe depression could be the cause of my symptoms when I called him on that he said he noted in his report ‘as tolerated’ that my chronic fatigue syndrome could also be the cause but he doesn’t deal with that only psychological causes. My therapist says I am clearly depressed because of my fatigue from what she has seen for a year. Being in constant pain, unable to participate in life day after day, month after month is depressing. Of course I feel useless. Of course I feel things might not get better. I haven’t been functioning for 16 months!”
>
> *Example 2*.
>
> “I could not sleep and went 15 days straight with no sleep. I was suicidal in addition to all the other Covid symptoms, thus one showed up two months after my acute Covid.”

Theme 19. Vibration and tremor symptoms caused disability for some people.

> *Example 1*.
>
> “I am writing to advise that I am one of the Covid long-haulers who is experiencing hand tremors. I feel them in my arms also and occasionally in my voice and breathing. The tremors in my hands are so severe that I cannot grip or hold a pen for any length of time before my hand writing deteriorates to chicken scratch. I have also resorted to dictating many of my emails and messages because my fingers don’t hit the right keys. I don’t know if tinnitus qualifies as internal vibration but I do have it and it is getting louder all the time. It causes me great anxiety and I have not been able to discover a treatment or solution.”
>
> *Example 2*.
>
> “Since March, I have had limited mobility as my legs give way and do not have the strength to walk unassisted. I have to use a chair to shower and walk with a cane. I cannot walk across the room without falling into things and struggle with balance. I have felt internal tremors that feel like a fizzing/bubbling that moves through my trunk and extremeties. My arms and legs shake and I have problems even with holding my fork still to feed myself. My mind doesn’t cooperate most days and I have speech issues with slurred speaking and stuttering.”

Theme 20. Vibrations and tremors could disturb or prevent sleep.

> *Example 1*.
>
> “Mostly Every morning waking up, there is an electrical zap from the top of my spine to mid back. Before I knew what the correct term was, I was telling doctors I buzz like a battery. This sensation happens first when I’m opening my eyes in the morning. It’s the first conscious feeling in the morning every day. If I try and go back to sleep the vibrations get more intense and more upsetting. So the best thing to do upon waking up is just get up and go on about my day. If I take a nap during the day. No problem, no vibrations. But there is a limit that I can sleep at any time, so if I do nap When I wake up, I don’t try and sleep more. There is I feel, a component of the vibrations That affect my sleep. I am very tired and feel most nights that my brains at war with itself and I don’t feel refreshed when waking up. Sometimes, not very often, I will get a whole body tremor feeling it’s unpleasant but, doesn’t last very long. I would say the ones that wake me up in the morning are more bothersome because I do not wake up gently or quietly. It really is internal torture.”
>
> *Example 2*.
>
> “Just when I thought I was done developing new symptoms in March this started, every time I start to fall asleep I get shooting pain and immense pressure in my arms, legs and spine. It wakes me up instantly. Imagine all the times you doze off a little in the day, times that by 20 if you are on medications that make you sleepy and try to imagine how torturous that symptom is when it happens to you 20 times a day. No one knows what this is or how to stop it. I’m forced to take muscle relaxers, lyrica, and Ativan to try to get to sleep before the symptom starts.”

Theme 21. Vibration and tremor symptoms could prevent people from working or carrying out daily life activities.

> *Example 1*.
>
> “I am not the person I was before Covid. I used to paint, refinish furniture, hang out with girlfriends, dance, golf, bike, travel, date. Now I barely have the energy to help my patient which is the only reason I have a roof over my head. If I can’t do it at any point, he needs to replace me and I lose my place to stay. I have no income and I couldn’t possibly work.”
>
> *Example 2*.
>
> “I got covid [September 2020]. I’m a nurse practitioner and cared for patients with Covid. I tried to go back to work. And after my psych Neuro testing I was found to have cognitive decline and severe memory recall and other memory issues. I was pulled from work [March], and then let go [May].”

Theme 22. Vibration and tremor symptoms could cause financial stress, through a combination of medical care costs and loss of income from medical leave.

> *Example 1*.
>
> “Before the pandemic was looking for another part time job so could live in a better place Now am drowning in medical debt with no relief in sight or ability to hold a job This has devastated my life Now am stuck living in an old garage without plumbing Hauling water back and forth from a garden hose and dumping dirty water takes what little energy have.”
>
> *Example 2*.
>
> “I got Covid [September 2020] and continue to have daily debilitating symptoms that have prevented me from going back to work. Side note: is there any government funding for those of us who are long haulers and can’t get back to work yet? My short term disability ran out beginning of April and I’ve been without any income since. It’s getting really tough.”

### Facebook

The 2 Word Clouds created revealed prevalent terms related to the themes found in emails. For all 450 Facebook comments combined, the 10 most frequent terms were tremors (64), covid (55), pain (51), vibrations (43), months (36), burning (29), feet (24), hands (22), legs (21), back (20).

The first Word Cloud analyzed 288 comments in response to a post that requested people experiencing tremors or internal vibrations to comment. This Word Cloud revealed that the five most common words used in comments were: tremors, covid, vibrations, months, and pain. Words included also indicated that symptoms ranged in presentation and severity, and vibrations were mentioned, as were seizures, shaking, and twitching. This Word Cloud also included other long Covid symptoms including brain fog, fatigue, and anxiety. Words indicating timing and duration of symptoms included months, days, and constantly. The only medication captured was gabapentin. Finally, similarly to the email responses, comments included mentions of sleep and tired.

**Figure 1.**
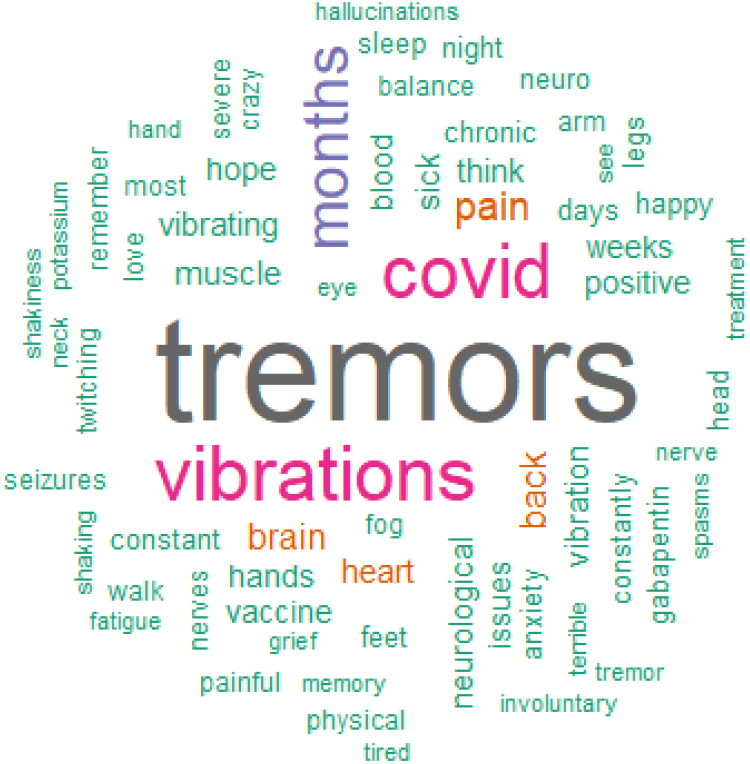
Word Cloud generated from Survivor Corps Facebook comments in response to post requesting information on tremor and internal vibrations.

The second Word Cloud was created based on 162 comments to a Survivor Corps poll that asked respondents about vibration or buzzing sensations and neuropathic pain. This Word Cloud revealed that pain, burning, covid, legs, hands, and feet were the most common terms mentioned in comments. This Word Cloud included terms not mentioned in the emails or the first Word Cloud, including sensations such as burning and symptoms such as shingles and thrush.

**Figure 2.**
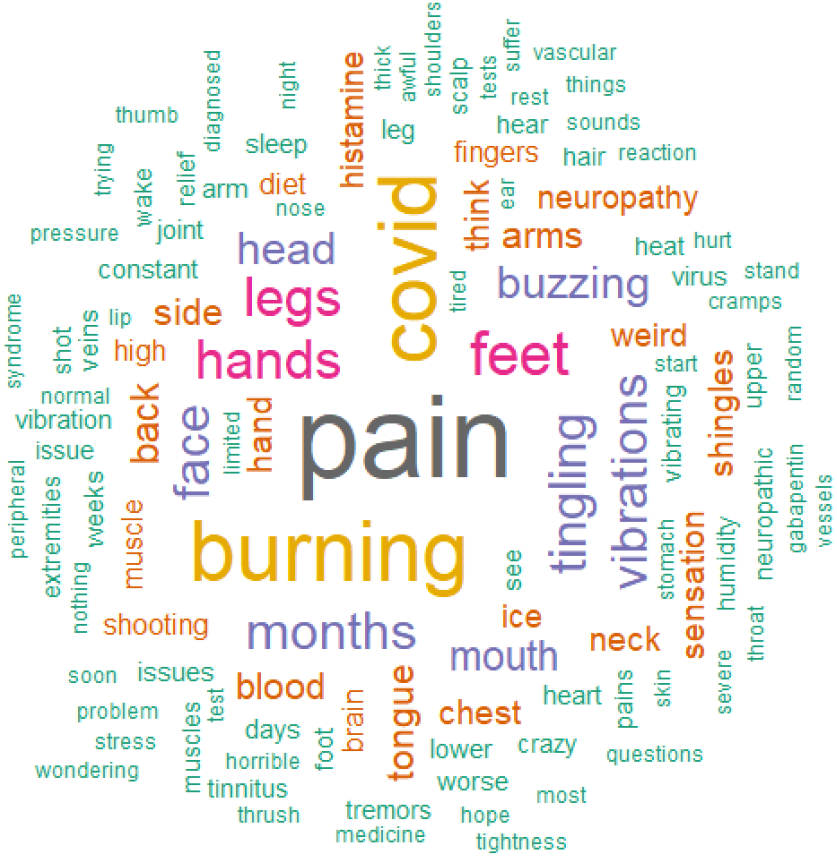
Word Cloud generated from Survivor Corps Facebook comments in response to poll related to vibrations, buzzing, and neuropathic pain.

## Discussion

These findings suggest that a group of people who report experiencing PASC exhibit a prolonged and debilitating symptom complex that prominently involves vibrations and tremors. While symptom experiences were heterogenous—in symptom timing, medical history, and initial infection, for instance—there were also common themes in how people described these symptoms and their effects. People also reported how testing and medical care have not yet identified possible mechanisms or successful treatment for these symptoms.

This study extends the literature in several ways. Previous literature consists of case series that provide preliminary reports of patients’ clinical presentation, course of care, and outcomes, but information on people’s experience with tremors more broadly has not been described in relation to long Covid.^5-12^ In addition, prior literature describes symptoms from the point of view of healthcare providers, but not from patients themselves. Previous case-series reports included a total of 16 people previously infected with SARS-CoV-2 who suffered from myoclonus-ataxia syndrome between 3 days to 6 weeks after acute, often mild or moderate, infection.^7-10,19-22^ Our report attempts to add a larger, broader overview of experiences of these symptoms, including symptoms described among individuals who might not have sought medical care.

This study, although limited in scope, is an effort to channel the perspective of patients for a condition that has yet to be defined. The utility of this study is that it may enable more formal and structured data collection for people with this syndrome. These experiences should be more rigorously characterized to develop hypotheses and understand mechanisms.

There are several key implications of the work. First, internal vibrations and tremors are causing severe suffering for a group of people after self-reported SARS-CoV-2 infection. Second, while the overall scale of these symptoms is still unknown, this group of people experiencing the symptoms have not recovered from the symptoms nor have they received specific diagnoses or been given treatment that completely alleviates their suffering. Third, the descriptions of feeling internal vibrations and tremor symptoms were similar across this group of patients. The emails were not seen by the other participants.

This study has limitations. The data included in this report are self-reported from people in response to a query. The information was limited to what people provided and there was no follow-up to obtain more information. The study presents what people shared on their initial communication. Also, some people did not have formal testing for infection. Any information about testing is based on self-report. In addition, there was little demographic information available for analysis. Also, because this is a convenience sample of people responding to the queries, this study cannot provide information on the incidence and prevalence of these symptoms.

In conclusion, some people report experiencing internal vibration and tremor symptoms, often causing intense suffering, after a self-reported history of SARS-CoV-2 infection. The symptoms had some common features but there was variability in timing, concomitant symptoms, and impact. Further research is needed to understand and alleviate this suffering, by studying the extent and scope of these symptoms, possible mechanisms, and potential treatment.

## Data Availability

All deidentified data in the present study are available upon reasonable request to the authors.

## Supplement 1

To solicit emails for this study, Survivor Corps and Nick Güthe requested emails from Survivor Corps members via Facebook and an emailed newsletter that is also posted on the Survivor Corps website.

First, Nick Güthe posted the following in the Survivor Corps Facebook page [July 2021]:

> *“Hi, to anyone on this group. It’s Nick Güthe, Heidi Ferrer’s husband. A study is forming with a top doctor for Long Haulers with Neurological tremors similar to Heidi Ferrer’s -- Tremors or internal vibrations. If you have these symptoms and want to be included please comment below. This isn’t a clinical trial but an attempt to gather data and stories to help get funding, bring attention to these symptoms which are so destructive to any Long Haulers physical and mental health*.*”*

Nick Güthe and Diana Zicklin Berrent responded to Facebook comments on this post to ask members to email their story, writing: “Send your story to.”

Second, a newsletter sent to members included the following [July 2021]:

> *“A study is forming for Long Haulers with Neurological tremors or internal vibrations. If you have these symptoms and want to be included, please* ***EMAIL*** *us with your details. This is not a clinical trial, but an attempt to gather data and stories to help get funding, and bring attention to these destructive symptoms that impact Long Hauler’s physical and mental health*.*”*

**Supplemental Table 1.**
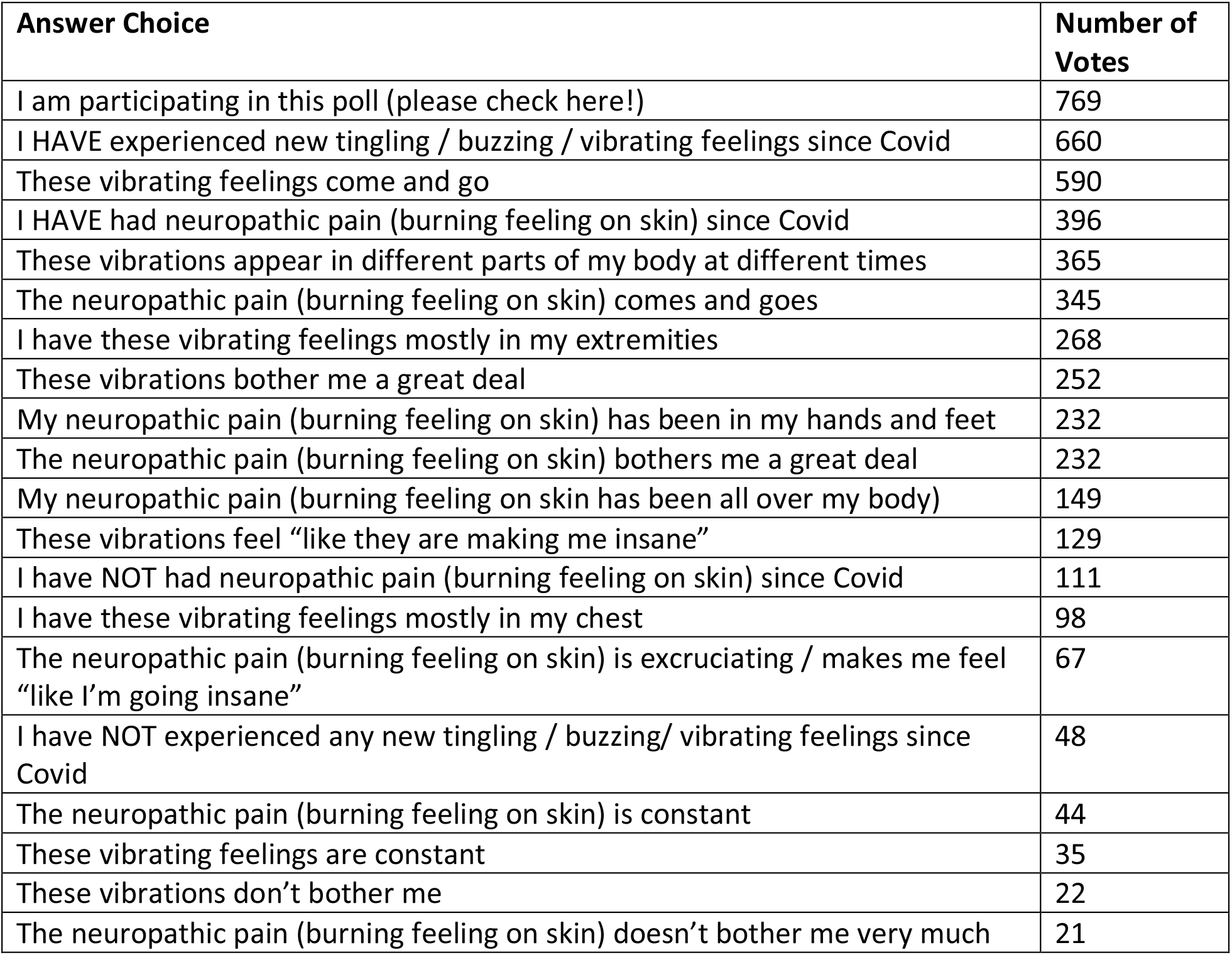
Full Survivor Corps Poll for Word Cloud 2. The following poll was posted in Survivor Corps [June 2021] and was titled “Vibration/Buzzing/Pain Poll.” There were 20 answer choices, each a statement relevant to vibration, tingling, buzzing, and neuropathic sensations. We have included the poll and responses for context, but for the purposes of qualitative analysis, we reviewed only the comments posted in response to this poll. These poll responses were collected as of July 16, 2021. The following text was included with the Facebook poll: *“VIBRATION / BUZZING / PAIN POLL* *SO MANY LONG HAULERS ARE DESCRIBING A BUZZING OR VIBRATION OR NEUROPATHIC PAIN IN THEIR BODIES AND EXTREMITIES - WE WANT TO LEARN MORE!”*

| Answer Choice | Number of Votes |
| --- | --- |
| I am participating in this poll (please check here!) | 769 |
| I HAVE experienced new tingling / buzzing / vibrating feelings since Covid | 660 |
| These vibrating feelings come and go | 590 |
| I HAVE had neuropathic pain (burning feeling on skin) since Covid | 396 |
| These vibrations appear in different parts of my body at different times | 365 |
| The neuropathic pain (burning feeling on skin) comes and goes | 345 |
| I have these vibrating feelings mostly in my extremities | 268 |
| These vibrations bother me a great deal | 252 |
| My neuropathic pain (burning feeling on skin) has been in my hands and feet | 232 |
| The neuropathic pain (burning feeling on skin) bothers me a great deal | 232 |
| My neuropathic pain (burning feeling on skin) has been all over my body | 149 |
| These vibrations feel “like they are making me insane” | 129 |
| I have NOT had neuropathic pain (burning feeling on skin) since Covid | 111 |
| I have these vibrating feelings mostly in my chest | 98 |
| The neuropathic pain (burning feeling on skin) is excruciating / makes me feel “like I’m going insane” | 67 |
| I have NOT experienced any new tingling / buzzing/ vibrating feelings since Covid | 48 |
| The neuropathic pain (burning feeling on skin) is constant | 44 |
| These vibrating feelings are constant | 35 |
| These vibrations don’t bother me | 22 |
| The neuropathic pain (burning feeling on skin) doesn’t bother me very much | 21 |

## References

1. Groff D, Sun A, Ssentongo AE, et al. Short-term and Long-term Rates of Postacute Sequelae of SARS-CoV-2 Infection: A Systematic Review. JAMA Network Open 2021;4:e2128568–e.

2. Davis HE, Assaf GS, McCorkell L, et al. Characterizing long COVID in an international cohort: 7 months of symptoms and their impact. EClinicalMedicine 2021;38.

3. Carfí A, Bernabei R, Landi F. Persistent Symptoms in Patients After Acute COVID-19. Jama 2020;324:603–5.

4. Huang C, Huang L, Wang Y, et al. 6-month consequences of COVID-19 in patients discharged from hospital: a cohort study. The Lancet 2021;397:220–32.

5. Przytuła F, Błądek S, Sławek J. Two COVID-19-related video-accompanied cases of severe ataxia-myoclonus syndrome. Neurol Neurochir Pol 2021;55:310–3.

6. Schellekens MMI, Bleeker-Rovers CP, Keurlings PAJ, Mummery CJ, Bloem BR. Reversible Myoclonus-Ataxia as a Postinfectious Manifestation of COVID-19. Movement disorders clinical practice 2020;7:977–9.

7. Emamikhah M, Babadi M, Mehrabani M, et al. Opsoclonus-myoclonus syndrome, a post-infectious neurologic complication of COVID-19: case series and review of literature. J Neurovirol 2021;27:26–34.

8. Shah PB, Desai SD. Opsoclonus Myoclonus Ataxia Syndrome in the Setting of COVID-19 Infection. Neurology 2021;96:33.

9. Dijkstra F, Van den Bossche T, Willekens B, Cras P, Crosiers D. Myoclonus and cerebellar ataxia following Coronavirus Disease 2019 (COVID-19). Mov Disord Clin Pract 2020;7:974–6.

10. Grimaldi S, Lagarde S, Harlé JR, Boucraut J, Guedj E. Autoimmune Encephalitis Concomitant with SARS-CoV-2 Infection: Insight from (18)F-FDG PET Imaging and Neuronal Autoantibodies. J Nucl Med 2020;61:1726–9.

11. Foucard C, San-Galli A, Tarrano C, Chaumont H, Lannuzel A, Roze E. Acute cerebellar ataxia and myoclonus with or without opsoclonus: a para-infectious syndrome associated with COVID-19. Eur J Neurol 2021;28:3533–6.

12. Wright D, Rowley R, Halks-Wellstead P, Anderson T, Wu TY. Abnormal Saccadic Oscillations Associated with Severe Acute Respiratory Syndrome Coronavirus 2 Encephalopathy and Ataxia. Movement disorders clinical practice 2020;7:980–2.

13. Massey D, Berrent D, Krumholz H. Breakthrough Symptomatic COVID-19 Infections Leading to Long Covid: Report from Long Covid Facebook Group Poll. medRxiv 2021:2021.07.23.21261030.

14. Lambert NSC. COVID-19 “Long Hauler” Symptoms Survey Report. Indian University School of Medicine 2020.

15. Lambert N, Corps S, El-Azab SA, et al. COVID-19 Survivors’ Reports of the Timing, Duration, and Health Impacts of Post-Acute Sequelae of SARS-CoV-2 (PASC) Infection. medRxiv 2021:2021.03.22.21254026.

16. Crabtree BF, Miller WL. Doing qualitative research. 2nd ed. Thousand Oaks, Calif.: Sage Publications; 1999.

17. Glaser BG, Strauss AL, ProQuest (Firm). The discovery of grounded theory strategies for qualitative research. New Brunswick (U.S.A): Aldine Transaction (a division of Transaction Publishers),; 1999:1 online resource.

18. Miles MB, Huberman AM. Qualitative data analysis : an expanded sourcebook. 2nd ed. Thousand Oaks: Sage Publications; 1994.

19. Przytuła F, Błądek S, Sławek J. Two COVID-19-related video-accompanied cases of severe ataxia-myoclonus syndrome. Neurologia i Neurochirurgia Polska 2021;55:310–3.

20. Schellekens MMI, Bleeker-Rovers CP, Keurlings PAJ, Mummery CJ, Bloem BR. Reversible Myoclonus-Ataxia as a Postinfectious Manifestation of COVID-19. Mov Disord Clin Pract 2020;7:977–9.

21. Foucard C, San-Galli A, Tarrano C, Chaumont H, Lannuzel A, Roze E. Acute cerebellar ataxia and myoclonus with or without opsoclonus: a parainfectious syndrome associated with COVID-19. Eur J Neurol 2021.

22. Wright D, Rowley R, Halks-Wellstead P, Anderson T, Wu TY. Abnormal Saccadic Oscillations Associated with Severe Acute Respiratory Syndrome Coronavirus 2 Encephalopathy and Ataxia. Mov Disord Clin Pract 2020;7:980–2.

